# The Canadian Pediatric Imaging Platform (C-PIP) Reference Sample: Study Design, Recruitment Strategy, and Phenotyping Protocol

**DOI:** 10.64898/2026.11.19.26368532

**Authors:** Samantha J. Lynch, Melissa Figueiredo, Filomeno Cortese, Mervyn Singh, Keith Owen Yeates, Miriam H. Beauchamp, Daniel C. Kopala Sibley, Donald J. Mabbott, Charles-Olivier Martin, Neta Bar Am, Dao Corriveau, Jordan Mak, Maria Mazaharally, Tomas Paus, Sean Spinney, Shireen Parimoo, Gregory A. Lodygensky, Helen L. Carlson, Ashley D. Harris, Lara M. Leijser, Kathryn Y. Manning, Jamie Near, G. Bruce Pike, Tamara Bodnar, Paul D. Arnold, Sébastien Jacquemont, Jacob A.S. Vorstman, Lune Bellec, Darren S. Kadis, Catherine Lebel, Sarah Lippé, Anne L. Wheeler, Signe Bray, Patricia Conrod

## Abstract

Distinguishing atypical from typical neurodevelopment requires large, well-characterized paediatric cohorts with harmonized multimodal neuroimaging and comprehensive phenotyping. Here, we describe the protocol for the Canadian Pediatric Imaging Platform (C-PIP) Reference Sample, including the design, recruitment strategy, and phenotyping protocol, along with overviews of the imaging and genetics protocols. Using a multisite accelerated longitudinal design, we aim to recruit >800 participants aged 0–16 years at baseline, across three Canadian sites (Alberta Children’s Hospital, Calgary; The Hospital for Sick Children (SickKids), Toronto; and CHU Sainte-Justine, Montreal) with comprehensive assessments at baseline and an annual follow-up, with additional follow-ups contingent on project extension. The protocol includes harmonized multimodal neuroimaging, extensive phenotyping across five broad domains: demographic, social and environmental factors; physical health and development; mental health and substance use; cognition; and temperament/personality, as well as DNA sequencing. Exclusion criteria are minimal to ensure a sample representative of the catchment areas of the participating hospitals. Data and protocols will be openly shared with the research community. The C-PIP Reference Sample will provide Canada’s first large-scale pediatric neuroimaging reference cohort, enabling researchers to map typical and atypical trajectories from infancy through adolescence, identify biomarkers, examine genetic and contextual influences on brain development, and provide a normative comparison framework for clinical populations.

## Introduction

Brain development is a complex and dynamic process, characterised by both linear and non-linear changes and substantial variation in the timing, pace, and pattern of maturation across individuals (Faghiri et al., 2019; Gozdas et al., 2018; Lebel et al., 2019; Mills et al., 2016; Mousley et al., 2025). Structural and functional changes occur across infancy, childhood and adolescence, and are influenced by interacting genetic, biological and environmental factors (Faghiri et al., 2019; Gozdas et al., 2018; Lebel et al., 2019; McLaughlin et al., 2019; Miguel et al., 2019; Mills et al., 2016; Zhou et al., 2023). Atypical developmental trajectories are associated with various forms of mental health and neurodevelopmental conditions, such as intellectual disabilities, autism spectrum disorder, attention deficit hyperactivity disorder, schizophrenia, anxiety and mood disorders (Appelbaum et al., 2022; Ball et al., 2017; Blok et al., 2022; Meyer C Lee, 2023). Understanding how and when developmental trajectories diverge may provide insight into the biological mechanisms underlying mental health and neurodevelopmental conditions and facilitate the identification of early markers of risk (Modabbernia et al., 2022; Muetzel et al., 2018; Parkes et al., 2021).

Large, longitudinal pediatric studies integrating multi-modal structural and functional neuroimaging, genetics and extensive phenotyping are needed to characterise normative developmental trajectories and establish reference models against which individual patterns of development can be compared (Becht C Mills, 2020; Cortese et al., 2023; Klapwijk et al., 2021; Zonneveld et al., 2020). Although several large-scale developmental neuroimaging initiatives have contributed to advancing our understanding of brain development (Baskin-Sommers et al., 2026; Morris et al., 2020; Paus et al., 2015; Schumann et al., 2010; Somerville et al., 2018), few longitudinal neuroimaging cohorts span the full developmental period from infancy through adolescence. As a result, normative developmental models are often derived from data covering restricted age ranges, limiting the ability to characterize trajectories across childhood and adolescence. Furthermore, variability in neuroimaging data acquisition platforms, imaging protocols, and cognitive, behavioural, and psychopathology measures across existing cohorts limits data integration and cross-study comparisons (Klapwijk et al., 2021; Zonneveld et al., 2020). Combined with the technical and logistical challenges associated with recruiting and retaining large, representative samples and acquiring high-quality imaging data in infants and younger children (Mills C Tamnes, 2014; Spann et al., 2023; Turesky et al., 2021), these barriers have hindered progress in mapping typical and atypical developmental trajectories. A cohort spanning infancy to late adolescence, therefore, offers a unique opportunity to establish normative reference trajectories across critical developmental transitions and to evaluate factors influencing neurodevelopment and mental health outcomes over time.

### The Canadian Pediatric Imaging Platform (C-PIP) Reference Sample

The Canadian Pediatric Imaging Platform (C-PIP) is a multisite, open science initiative developed to address the above challenges and accelerate discovery in pediatric brain health. A central goal of this initiative is to establish Canada’s first longitudinal reference sample of >800 children aged 0-16 years. Using an accelerated longitudinal design, this reference cohort is designed to provide normative benchmarks for brain development and associated psychopathology, cognitive, behavioural, and environmental factors. The protocol is designed to be sensitive to the identification of atypical developmental trajectories and brain-based markers of neurodevelopmental and mental health risk and includes measures relevant to key exposures and clinical populations such as preterm birth, prenatal exposure to substances, acquired brain injury, genetic syndromes, and early adverse experiences. Additionally, many neuroimaging and phenotyping measures align with those used in major international developmental cohorts, facilitating cross-cohort comparisons, replication, and future data integration efforts. Data will be made available to qualified users in a series of curated releases to support reproducibility, scalability, and integration with clinical and translational research. Protocols and processing pipelines will be openly available via GitHub (https://bids-ffux-docs.readthedocs.io/en/latest/). Here we describe the C-PIP reference sample study, including the design, recruitment strategy, phenotyping protocol, and genetics protocol. The neuroimaging protocol is described in detail in an accompanying manuscript (Singh et al., under review).

### C-PIP Reference Sample Study Design

#### Participants

We aim to recruit 816 participants from 0 to 16 years of age at three sites: Alberta Children’s Hospital in Calgary (University of Calgary), SickKids Hospital in Toronto (University of Toronto) and Centre Hospitalier Ste-Justine (CHUSJ, University of Montreal). Recruitment commenced in January 2025. As shown in Figure 1, recruitment will prioritize younger children (0 to 6 years) to account for the higher likelihood of incomplete or failed scans in this group.

**Figure 1.**
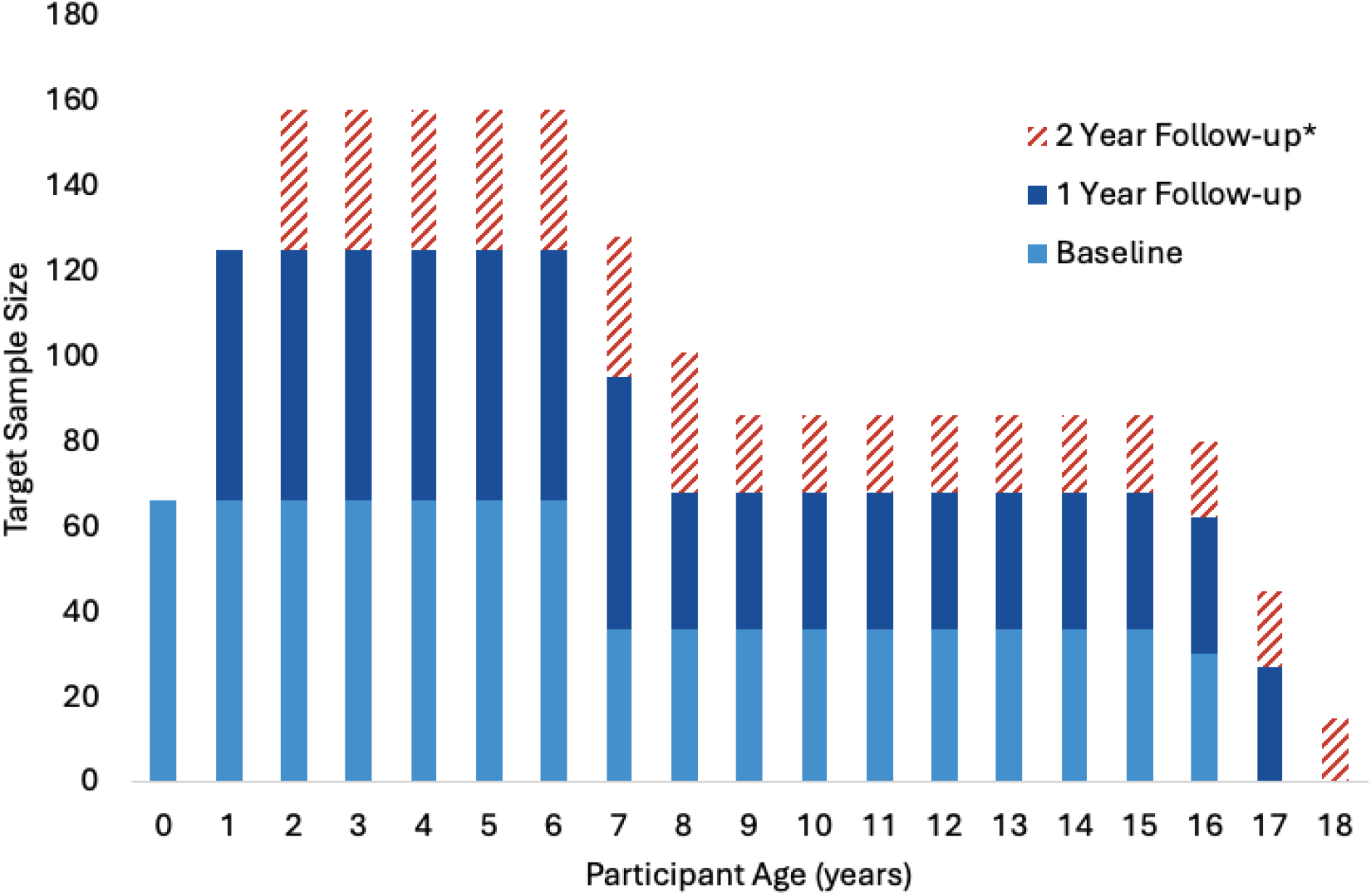
C-PIP Recruitment and Retention Targets by Age. **Note.** *2 Year Follow-up is contingent on project extensions. Target retention rates are 90% at 1 Year Follow-up, and 50% at 2 Year Follow-up.

C-PIP aims to recruit a sample that is representative of children of the catchment area of these three large pediatric hospitals, based on available census data. Recruitment targets for sex, ethnicity/race, socioeconomic (mid-high / low SES) and geographic (urban/rural) characteristics were derived from census data for the catchment areas of the three sites (Table 1) and are monitored regularly to allow sites to adjust recruitment tactics as needed.

**Table 1.**
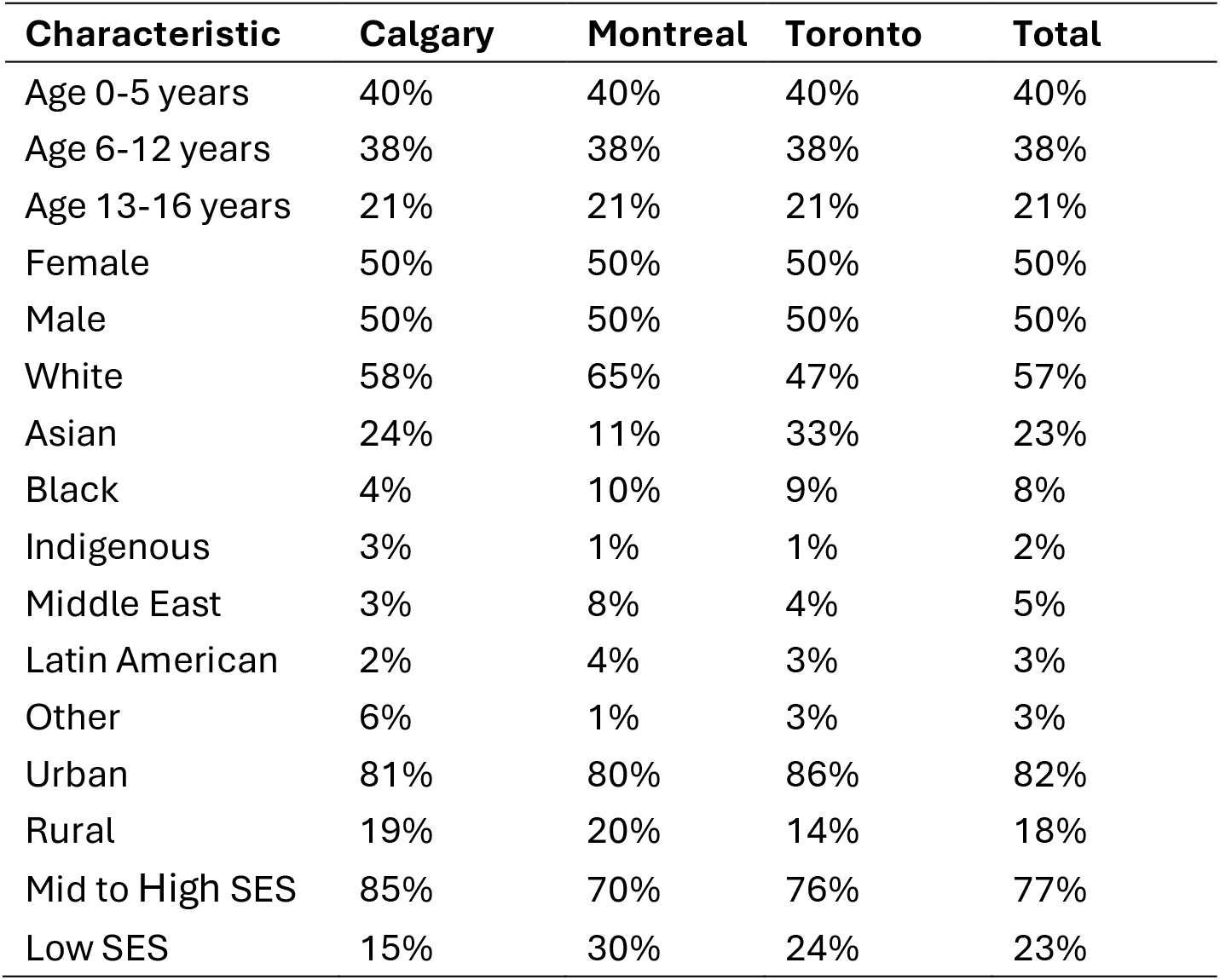
Target recruitment distribution for the C-PIP reference sample derived from census estimates for the catchment areas of the participating sites.

| **Characteristic** | **Calgary** | **Montreal** | **Toronto** | **Total** |
|---|---|---|---|---|
| Age 0-5 years | 40% | 40% | 40% | 40% |
| Age 6-12 years | 38% | 38% | 38% | 38% |
| Age 13-16 years | 21% | 21% | 21% | 21% |
| Female | 50% | 50% | 50% | 50% |
| Male | 50% | 50% | 50% | 50% |
| White | 58% | 65% | 47% | 57% |
| Asian | 24% | 11% | 33% | 23% |
| Black | 4% | 10% | 9% | 8% |
| Indigenous | 3% | 1% | 1% | 2% |
| Middle East | 3% | 8% | 4% | 5% |
| Latin American | 2% | 4% | 3% | 3% |
| Other | 6% | 1% | 3% | 3% |
| Urban | 81% | 80% | 86% | 82% |
| Rural | 19% | 20% | 14% | 18% |
| Mid to High SES | 85% | 70% | 76% | 77% |
| Low SES | 15% | 30% | 24% | 23% |

#### Inclusion and exclusion criteria

Inclusion criteria are that children must be 0–16.99 years of age at baseline (up to, but not including, their 17^th^ birthday). Where possible, accommodations will be made for families with language needs beyond English or French (e.g., interpreter services).

To ensure that the reference sample is phenotypically diverse and representative of the general Canadian population, exclusion criteria are minimal and include 1) MRI contraindications (e.g., braces, implantable medical devices, inability to undergo an MRI without sedation), and 2) inability or unwillingness to provide informed consent including consent to re-use MRI and phenotypic data by other research teams. While these criteria are necessary for participant safety and data quality, they may limit representation of children with severe neurodevelopmental impairments or behavioural difficulties.

#### Ethical approvals

Local advisory committees including parents and clinicians informed the development of the study protocol, and a multidisciplinary ethics and data governance working group was established to ensure that ethics approvals were obtained and language around data sharing was consistent across sites. The study was approved by research ethics boards at each site (ACH: REB22-1749, CHUSJ: 2024-6249, SickKids: 2984).

#### Recruitment

Participants will be recruited within each of the participating hospitals, which are major pediatric referral centres, allowing for the recruitment of a socioeconomically and demographically representative sample. Emergency Departments (EDs) and hospital registries, where possible, will serve as the primary recruitment source. This takes advantage of the normative nature of ED attendance among Canadian children (Canadian Institute for Health Information, 2024), where visits are often prompted by acute illnesses or injuries (e.g., asthma, upper respiratory infections, head injuries) (Seguin et al., 2018; Tiller et al., 2021). Each hospital has a pediatric emergency department research recruitment program, through which volunteers, research assistants, medical students, or research nurses approach families, provide a brief description of the study, and invite families to provide consent-to-contact (ACH, CHUSJ) or to enroll in the study (SickKids). The Montreal (CHUSJ) site will additionally leverage a database of all children born at CHUSJ. Prospective participants are identified within the database based on the study inclusion/exclusion criteria. The research team send an invitation to prospective participants by mail in the first instance and then email or phone depending on response to the mailing. Sites may also supplement recruitment with advertising throughout their hospital or through targeted social media, postpartum unit, and online advertising or other community outreach initiatives to enhance representativeness and recruitment efficiency.

#### Study ffow

An overview of the study flow, from recruitment through annual follow-ups, is shown in Figure 2. Families who are not consented in the ED will be contacted over the phone or invited to attend a virtual meeting where the study is explained in detail and informed consent/assent is obtained (via eConsent). During this session, families are screened for MRI contraindications, and the first in-person site visit is scheduled.

**Figure 2.**
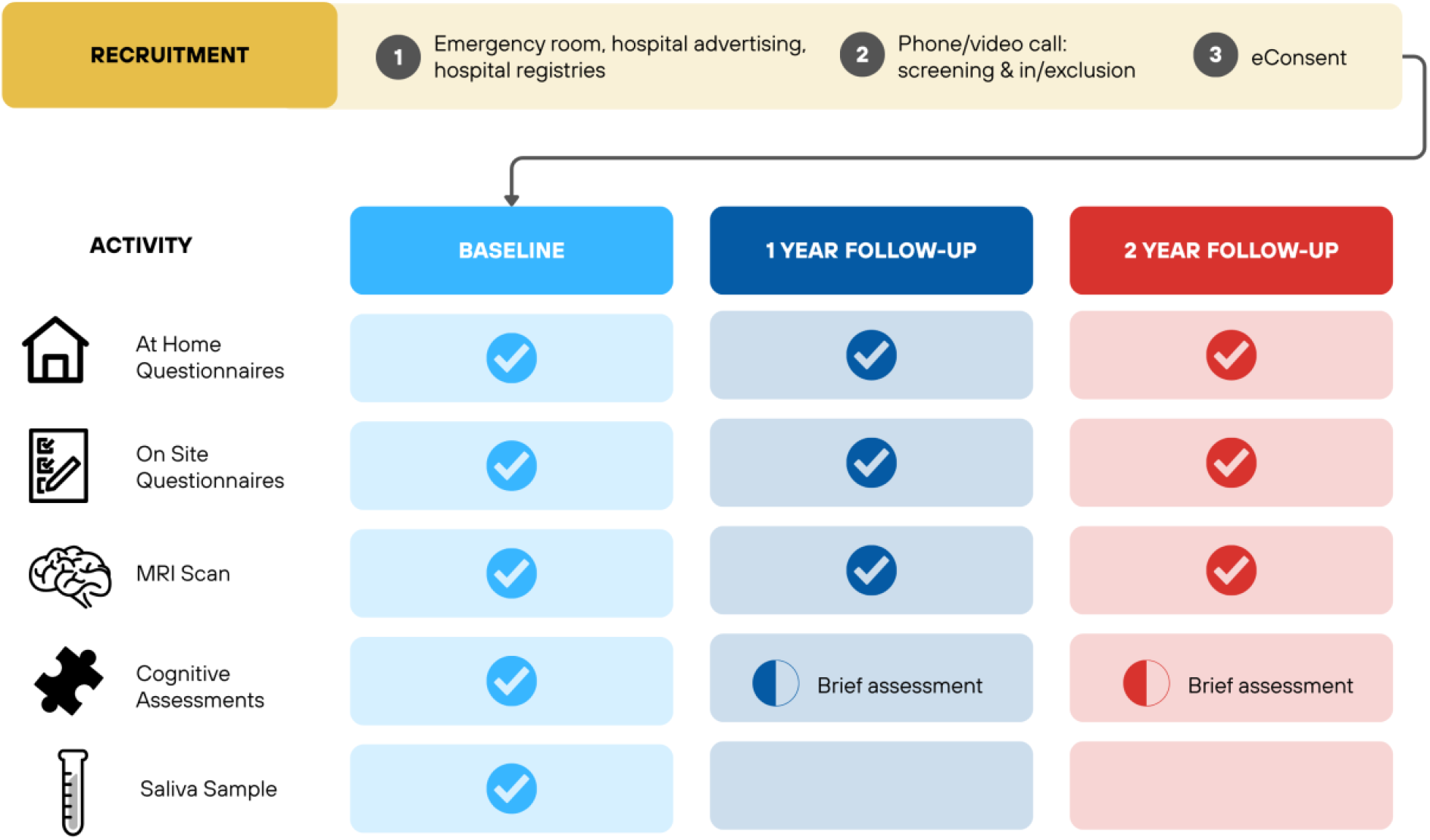
C-PIP Reference Sample Cohort Study Flow.

Participants then complete a baseline site visit, with planned follow-up approximately 1 year post-baseline. Follow-up intervals may be shortened to maximize longitudinal data collection within the study period. Additional follow-ups are planned through project extensions.

Each site visit includes an MRI scan, standardized age-appropriate cognitive assessment, and questionnaires. To reduce burden, caregivers and youth aged 11+ years are sent questionnaires to complete at home in the two weeks prior to their visit. The cognitive assessment is shortened at follow-up, and a saliva sample for DNA sequencing is collected at baseline only.

#### Site visit overview

On the day of the site visit, children complete an MRI scan, cognitive assessments, and questionnaires as outlined in the Neuroimaging and Phenotyping Protocol sections below. At baseline, a saliva sample is collected for genotyping (further details below). Caregivers complete the on-site questionnaires while the child is being scanned and/or completing the cognitive assessment. Families have the opportunity for an extended break between the cognitive assessment and the MRI portion of the visit. The MRI is typically scheduled after phenotyping assessments to capitalise on participant fatigue and reduce risk of movement during scanning. However, the order can be adapted, or split across two days, to best accommodate families and optimize data quality and participant retention.

### Neuroimaging protocol

C-PIP has implemented a multimodal neuroimaging protocol to comprehensively characterise brain structure and function. Detailed information on the neuroimaging protocol is reported in a separate publication (Singh et al., ref). Briefly, the C-PIP protocol includes the following core sequences: (1) a high-quality T1- and T2-weighted structural imaging (prioritised according to the age of the child), (2) multi-shell diffusion imaging (Farquharson C Tournier, 2015), and (3) movie-watching multi-echo functional imaging (Kundu et al., 2017). Additional sequences, including (4) magnetisation prepared 2-rapid gradient echo imaging, (5) inhomogeneous magnetisation transfer imaging (Varma et al., 2015); (6) strategically acquired gradient echo imaging (Chen et al., 2018), and (7) whole-slice magnetic resonance spectroscopic imaging (Zhu C Barker, 2011) were incorporated to probe other aspects of brain composition. All sequences were designed to be short as possible to maximise the feasibility of obtaining high-quality data, with optimised parameters to further improve compliance and sensitivity to tissue contrast-related differences in infants and toddlers (< 3 years).

### Phenotyping Protocol

#### Assessment Domains

Participants and their caregivers complete questionnaires and standardized direct assessments covering five broad domains: demographic, social and environmental factors; physical health and development; mental health and substance use; cognition; and temperament/personality. Drawing on recent large scale developmental neuroimaging cohort studies (Barch et al., 2018; Schumann et al., 2010; Somerville et al., 2018), the selection of measures was guided by: 1) alignment with existing literature on relevant and developmentally appropriate constructs; 2) applicability across different developmental stages (or availability of parallel versions across stages); 3) validation in both English and French, and 4) sensitivity to change over time. The selected domains and constructs enable a comprehensive phenotyping of factors related to brain development from infancy through adolescence. An overview of assessment domains by age group is provided in Table 2, and briefly described below. Detailed information about the measures across child age and timepoint provided in the Supplementary Materials.

**Table 2.** Summary of phenotyping domains and measures included in the C-PIP reference sample.

| Domain/Construct/Measure | Source/Reporter | Time to complete | **Infant** (0–2yrs) | **Child** (3–10yrs) | **Adolescent** (11+yrs) |
|---|---|---|---|---|---|
| **Demographic, social & environmental factors** |  |  |  |  |  |
| Demographics & household factors | C & Y | 3 mins | X | X | X |
| Sexual orientation & romantic relationship history | Y | 1 min | – | – | X |
| Adverse life events | C & Y | 3 mins | X | X | X |
| Digital media use | C & Y | 2 mins | X | X | X |
| Family psychiatric history | C | 4 mins | X | X | X |
| Caregiver mental health/psychological distress | C | 1 min | X | X | X |
| Caregiver substance use | C | 5 mins | X | X | X |
| Caregiver personality | C | 5 mins | X | X | X |
| Caregiver digital media use | C | 1 min | X | X | X |
| **Physical health, lifestyle & development** |  |  |  |  |  |
| Developmental history, Pregnancy & Birth Questionnaire | C | 10 mins | X | – | – |
| Medical History Questionnaire | C | 5 mins | X | X | X |
| Concussion & symptoms | C & Y | 3 mins | X | X | X |
| Pubertal development | C & Y | 1 min | – | X | X |
| Exercise | Y | 1 min | – | – | X |
| Sleep disturbances & impairments | C & Y | 1 min | X | X | X |
| **Mental health & substance use** |  |  |  |  |  |
| Dimensional psychopathology (P-CBCL, CBCL, YSR) | C & Y | 10-20 mins | X | X | X |
| Eating, psychosis & social phobia screening | Y | 4 mins | – | – | X |
| Substance use & harms | Y | 4 mins | – | – | X |
| **Temperament & personality** |  |  |  |  |  |
| Temperament/personality | C & Y/R | 4-7 mins | X | X | X |
| Substance use personality risk profiles | C & Y/R | 4 mins | – | X | X |
| **Cognition** |  |  |  |  |  |
| Cognitive assessment (Bayley-IV, WPPSI-IV, WISC-V, WAIS-IV) | R | 60-90 mins | X | X | X |
| Cognitive flexibility (NIH Dimensional Change Card Sort) | R | 10 mins | – | X | X |
| Inhibitory control (Bear/Dragon, NIH Flanker) | R | 15 mins | X | X | X |

Note. Medical and family psychiatric history questionnaires are administered at baseline and T3. C = Caregiver, Y = Youth (predominantly adolescents, some self-report instruments are administered to 8-10 yr olds on site), R = Research Assistant. Developmental history (pregnancy, birth and early development questions) assessed at baseline only. Bayley-IV = Bayley Scales of Infant and Toddler Development, Fourth Edition, CBCL = Child Behavior Checklist, P-CBCL = Preschool Child Behavior Checklist , WPPSI-IV = Wechsler Preschool and Primary Scale of Intelligence, Fourth Edition: Canadian, WISC-IV = Wechsler Intelligence Scale for Children, Fifth Edition: Canadian, WAIS-IV = Wechsler Adult Intelligence Scale, Fourth Edition: Canadian, YSR = Youth Self Report.

##### Demographic, social & environmental factors

Measures of demographic, social and environmental factors were selected to characterise developmental contexts, including family structure, adverse experiences, and contemporary exposures, such as digital media use for both the participant and the caregiver (Baskin-Sommers et al., 2026; Cai et al., 2021; Ferschmann et al., 2022a; Holz et al., 2023; Kessler et al., 2010; Malave et al., 2022; Miguel et al., 2019; Patterson C Vakili, 2014; Tooley et al., 2021; Whittle et al., 2024). Additionally, questionnaires about the caregivers substance use, psychological distress, personality, and digital media use were included in recognition of the role that these factors play in contributing to the developmental context of a child, and brain and mental health outcomes (Aguinaldo et al., 2026; Ferschmann et al., 2022b; Kessler et al., 2010; Kwarteng et al., 2021; Schofield et al., 2012).

##### Physical health, lifestyle and development

Measures spanning pregnancy/birth complications, developmental history, medical history, concussion, puberty, exercise, sleep were selected based on established links to brain health and maturational processes (Beauchamp et al., 2020; Dennis, Caeyenberghs, Asarnow, et al., 2021; Dennis, Caeyenberghs, Hoskinson, et al., 2021; Giesbrecht et al., 2022; Lebel et al., 2012; Lindström et al., 2011; Long C Lebel, 2022; Mandl et al., 2024; Nguyen et al., 2019; Shukla et al., 2021; Stojanovski et al., 2019, 2021; Ware et al., 2022; Yeates et al., 2017).

##### Mental health and substance use

Dimensional measures of psychopathology, rather than categorical diagnostics, were selected based on evidence supporting dimensional approaches for identifying brain-based markers of atypical development (Modabbernia et al., 2022; Muetzel et al., 2018; Parkes et al., 2021). These approaches also facilitate research aligned with contemporary frameworks, such the Research Domain Criteria (RDoC)(Cuthbert, 2022; Insel et al., 2010) and the Hierarchical Taxonomy of Psychopathology (HiTOP) (Kotov et al., 2017). Adolescents (11+ years) also complete measures assessing lifetime and recent substance use patterns, substance-related harms, and are screened for alcohol and cannabis use disorders.

##### Temperament and personality

Age-appropriate measures of temperament and personality were selected to capture individual differences in dispositional tendencies across development (Beaty et al., 2016; Conrod C Nikolaou, 2016a; Ferschmann et al., 2018; Zhang et al., 2023). Temperament measures administered in infancy and childhood assess broad domains of negative emotionality/affect, surgency or positive affectivity, and self-regulatory capacity (effortful control/orienting-regulation), whereas personality measures administered in later childhood and adolescence assess the Big Five domains of extraversion, agreeableness, conscientiousness, negative emotionality, and open-mindedness. Using measures with conceptual continuity enables modelling of trait stability and change, and of the transition from infant temperament to Big Five personality, alongside brain development. Adolescents (11+ years) and older children (8-10 yrs) also complete the Substance Use Risk Profile Scale (SURPS), which assesses four personality traits associated with elevated risk for substance use and co-occurring mental health problems: hopelessness, anxiety sensitivity, impulsivity, and sensation seeking (Conrod C Nikolaou, 2016b; Woicik et al., 2009).

#### Cognition

Cognitive measures provide objective, task-based indices of typical maturation and neurodevelopmental variation (Anokhin et al., 2022). Age-appropriate standardized scales, including the Bayley Scales of Infant and Toddler Development, Fourth Edition (BSID-4) (Bayley C Aylward, 2021), the Wechsler Preschool and Primary Scale of Intelligence, Fourth Edition (WPPSI-IV) (Wechsler, 2012, 2013), the Wechsler Intelligence Scale for Children, Fifth Edition (WISC-V) (Wechsler, 2015a, 2015b) and the Wechsler Adult Intelligence Scale, Fourth Edition: Canadian (Wechsler, 2008) (Wechsler, 2012, 2015a)(Bayley C Aylward, 2021)were selected for their comprehensive domain coverage, sensitivity to developmental change and deviations, and widespread clinical use, enhancing translational potential. NIH Toolbox tasks (Flanker and Dimensional Change Card Sort) were included to support harmonization with large-scale cohorts and provide brief, validated executive function measures suitable for annual administration (Anokhin et al., 2022; Weintraub et al., 2013; Zelazo et al., 2013).

A small number of questionnaires, primarily assessing demographic and physical health, lifestyle, and development factors (e.g., medical history, exercise, concussion) did not have existing versions available in both French and English. The specific questionnaires requiring translation are identified in the Supplementary Materials (Table S1). Where required, questionnaires were translated into the target language by a bilingual member of the research team and reviewed by a second bilingual team member. Permission was sought from the developer or copyright holder when necessary. For all but one questionnaire, the translation was from English to French.

#### Assessment administration

Most assessments at both baseline and follow-up, except for domains, are unlikely to change (e.g., birth/pregnancy and early childhood development). Caregivers are the primary reporters in early childhood, with youth self-report introduced from 11 years of age (note. the Health Behaviour Inventory and personality measures for 8-10 yr olds are also administered as self-report measures; these are completed in person in the presence of a research assistant who can provide support if needed). Assessments are completed on-site with measures administered by trained research staff (1.5-2 hours). Median questionnaire completion time is approximately 45–60 minutes for youth (7–15 minutes at home and 25–45 minutes on-site) and approximately 1.5 hours for caregivers after adjusting for idle time (30–50 minutes on-site and 40–195 minutes at home).

To ensure appropriateness and acceptability of the measures, the protocol was reviewed by two external advisory committees, one comprised of parents and one comprised of external researchers. In addition, the protocol was piloted with children and their parents across all three sites (Total N = 12: 0-2 yrs n = 1; 3-6 yrs n = 4; 7-10 yrs n = 4; 11-16 yrs n = 3).

#### REDCap database design and development

Questionnaires and other non-imaging data (e.g., cognitive assessment forms, MRI session log) will be collected using REDCap, – a secure, web-based program that is widely supported by research institutions (Harris et al., 2009, 2019). As shown in Figure 3, two separate REDCap databases were developed to support administrative and research functions. A customisable ***Admin* database** template was created to manage participant screening, eConsent, contact information, and visit planning. It includes core screening and eligibility forms, as well as example eConsent templates. This template was uploaded to each site’s local REDCap server and adapted to meet local requirements (e.g., site-specific consent forms).

**Figure 3.**
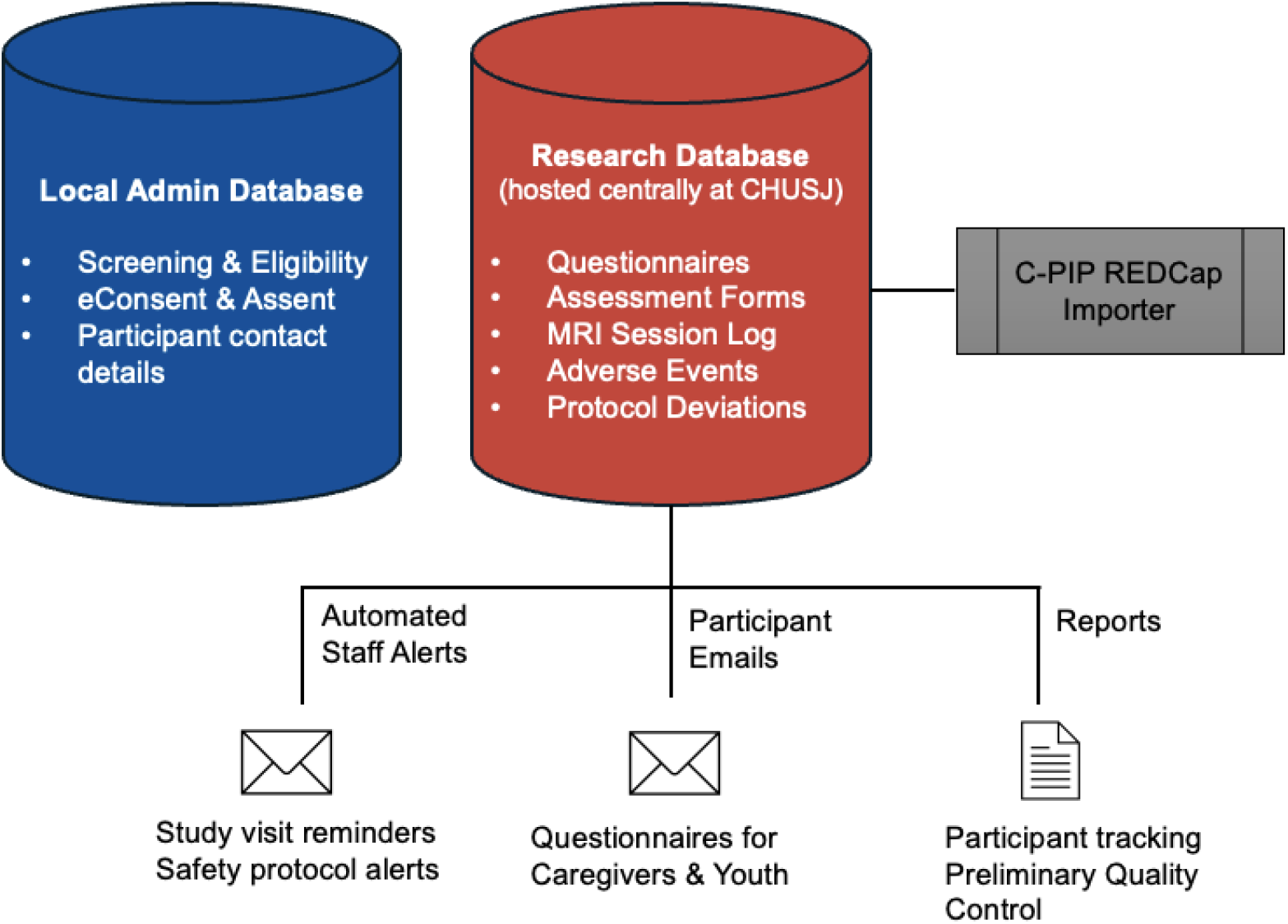
Overview of REDCap Database Design.

A centralised ***Research* database** (hosted at CHUSJ) was designed to collect all non-imaging study data, including responses to questionnaires, staff/examiner forms, MRI session logs, and other research logs (e.g., adverse events, protocol deviations, study withdrawals). To implement the age-dependent assessment protocol, questionnaires were first programmed individually and then grouped into five core dynamic forms:

1 Caregiver – At Home Questionnaires
2) – At Home Questionnaires
3) Caregiver – On Site Questionnaires
4) Youth – On Site Questionnaires
5) Staff – All Questionnaires C Tasks

These dynamic forms allow tailored administration based on participant age and measurement occasion. At-home forms can be emailed for completion on participants’ personal devices, while on-site forms can be securely administered via study iPads. The multi-language module enables participants to complete questionnaires in their preferred language. English and French versions of all questionnaires are currently available, and other languages can be easily added, enhancing global utility. Staff forms enable research staff to track completion of all required assessments during the site visit. The *Admin* and *Research* REDCap databases were developed with future scalability in mind and can be shared with other research teams to support data collection at additional sites or in related studies. Further details about the integration of data collected via REDCap with neuroimaging and genetic data will be described elsewhere (Spinney et al., under review).

### Genetics protocol

Saliva samples will be collected using Oragene-DNA kits (OG-675 for children and adolescents, and OC-175 for infants and children up to 5 years old if necessary). Genotyping will be performed using the Illumina Infinium Global Screening Array-24, which is widely used in genome-wide association studies (GWAS) studies to detect common genetic variants (Van Asselt C Ehli, 2022). Polygenic risk scores (PRS) will be calculated to examine associations between aggregated genetic risk and brain development. Data will be made available via the C-PIP data sharing infrastructure to approved users.

### Governance

C-PIP is governed by an Executive Committee and a Scientific Committee, which includes the site leads, site coordinators, and Working Group leads. Five cross-site Working Groups oversee phenotyping and reference sample study implementation (Leads: Conrod, Lynch, Lippe), neuroimaging (Lead: Wheeler), IT/data management (Lead: Bellec), equity diversity, inclusion, and accessibility (EDIA; Lead: Bray), and ethics/data governance (Lead: Lebel). This structure supports coordinated decision-making, protocol harmonization, quality control, and implementation across participating sites.

### Discussion: Anticipated Outcomes and Opportunities

The C-PIP reference sample cohort and associated platform are poised to catalyze progress in understanding pediatric neurodevelopment from birth through adolescence. The primary outcome of the reference sample will be the characterisation of normative developmental trajectories that can serve as benchmarks for identifying atypical development through comparison with clinical samples. By combining comprehensive phenotyping assessments, high-quality, multimodal neuroimaging, genetics, and an accelerated longitudinal design, C-PIP will provide a unique resource for examining how individual and contextual factors influence brain development and mental health outcomes across critical periods. This rich, multimodal dataset will enable researchers to map typical and atypical developmental trajectories, identify early biomarkers associated with diverse outcomes, and provide a normative comparison framework for clinical populations. As Canada’s first large-scale pediatric neuroimaging reference cohort, C-PIP will support robust case-control trajectory modelling for a variety of functional outcomes relevant to mental health, neurodevelopmental conditions, and traumatic brain injuries. In parallel with the development of the reference sample, two clinical demonstration projects were established, one focusing on concussion and the other focussing on selected neurodevelopmental conditions and genetic profiles. These projects will illustrate how the reference cohort can be leveraged to address condition-specific research questions and will be described in subsequent papers. In summary, the C-PIP reference sample cohort will provide a foundational resource for a new generation of pediatric neuroscience research in Canada and internationally, enabling characterisation of individual neurodevelopmental variation and supporting translational research across diverse clinical applications.

## Data Availability

All data produced in the present study are available upon reasonable request to the authors

## CRediT authorship contribution statement

**Samantha J. Lynch**: Conceptualization, Data curation, Project administration, Visualisation, Writing – original draft, Writing – review C editing. **Melissa Figueiredo**: Methodology, Investigation, Visualisation, Writing – review C editing. **Keith Yeates**: Conceptualization, Methodology, Funding acquisition, Writing – review C editing. **Miriam Beauchamp**: Conceptualization, Methodology, Resources, Funding acquisition, Writing – review C editing. **Daniel C. Kopala-Sibley**: Conceptualization, Methodology, Writing – review C editing. **Donald Mabbott**: Conceptualization, Methodology, Funding acquisition, Writing – review C editing. **Charles-Olivier Martin**: Methodology, Investigation, Resources, Supervision, Writing – review C editing. **Filomeno Cortese**: Methodology, Project administration, Supervision, Writing – review C editing. **Neta Bar Am**: Investigation, Project administration, Writing – review C editing. **Dao Corriveau**: Investigation, Project administration, Writing – review C editing. **Jordan Mak**: Investigation, Project administration, Writing – review C editing. **Maria Mazaharally**: Investigation, Project administration, Writing – review C editing. **Sébastien Jacquemont**: Conceptualization, Methodology, Funding acquisition, Writing – review C editing. **Tomas Paus**: Conceptualization, Methodology, Resources, Funding acquisition, Writing – review C editing. **Mervyn Singh**: Conceptualization, Methodology, Project administration, Writing – review C editing. **Shireen Parimoo**: Conceptualization, Methodology, Writing – review C editing. **Sean Spinney**: Methodology, Data curation, Software, Writing – review C editing. **Gregory A. Lodygensky**: Conceptualization, Methodology, Funding acquisition, Writing – review C editing. **Helen Carlson**: Conceptualization, Methodology, Writing – review C editing. **Ashley D. Harris**: Conceptualization, Methodology, Funding acquisition, Writing – review C editing. **Lara M. Leijser**: Conceptualization, Methodology, Writing – review C editing. **Kathryn Manning**: Conceptualization, Methodology, Writing – review C editing. **Jamie Near**: Conceptualization, Methodology, Funding acquisition, Writing – review C editing. **Bruce Pike**: Conceptualization, Methodology, Funding acquisition, Writing – review C editing. **Paul Arnold**: Conceptualization, Methodology, Funding acquisition, Writing – review C editing. **Jacob Vorstman**: Conceptualization, Methodology, Writing – review C editing. **Tamara Bodnar**: Conceptualization, Methodology, Writing – review C editing. **Lune Bellec**: Conceptualization, Methodology, Data curation, Software, Supervision, Funding acquisition, Writing – review C editing. **Darren Kadis**: Conceptualization, Methodology, Funding acquisition, Writing – review C editing. **Catherine Lebel**: Conceptualization, Methodology, Supervision, Funding acquisition, Writing – review C editing. **Sarah Lippé**: Conceptualization, Methodology, Resources, Supervision, Writing – review C editing. **Anne L. Wheeler**: Conceptualization, Methodology, Supervision, Funding acquisition, Writing – review C editing. **Signe Bray**: Conceptualization, Methodology, Supervision, Funding acquisition, Writing – original draft, Writing – review C editing. **Patricia Conrod**: Conceptualization, Methodology, Supervision, Funding acquisition, Writing – original draft, Writing – review C editing.

## Acknowledgements

We gratefully acknowledge the contributions of the research staff for their outstanding efforts towards participant recruitment and data collection, including Charlotte Bibeau, Angelina Haddad, Frédérique Marcoux, Karen Miller-Zajdman, Michelle Nehme, Olatioluwase Olatona, Sophia Rivard, Marion Royer D’Halluin, and Joshua Valcourt. We also thank Janelle Zhan (Unité de recherche clinique appliquée (URCA), Direction de la recherche, CHU Sainte-Justine), for her invaluable support with REDCap programming and database management. Finally, we extend our sincere thanks to the families who generously participated in piloting the study protocols and provided valuable feedback that helped inform the study procedures.

## Declaration of generative AI and AI-assisted technologies in the manuscript preparation process

Microsoft CoPilot and OpenAI ChatGPT (GPT-4 and GPT-5 series) were used to assist with editing and refinement of text in the manuscript, including condensing wording. After using these tools, the authors reviewed and edited content as needed. The authors take full responsibility for the content of the manuscript.

