## Supplementary Materials for "The Canadian Pediatric Imaging Platform (C-PIP) Reference Sample: Study Design, Recruitment Strategy, and Phenotyping Protocol"

#### Table of Contents

Table S1. C-PIP Phenotyping Assessment Protocol by Age of Child and Measurement Occasion . 2

Table S2. Baseline Demographics ........................................................................................ 6

Table S3. Wechsler Scale Subtests Order of Administration and Composite Scores .................. 7

#### Corresponding Authors

**Samantha J. Lynch**
Azrieli Research Center, CHU Sainte-Justine Mother–Child University Hospital
3175 Côte-Sainte-Catherine, Montréal, QC H3T 1C5, Canada


**Patricia Conrod**
Azrieli Research Center, CHU Sainte-Justine Mother–Child University Hospital
3175 Côte-Sainte-Catherine, Montréal, QC H3T 1C5, Canada


**Signe Bray**
Child and Adolescent Imaging Research (CAIR) Program, Alberta Children's Research Institute, University of Calgary, 28 Oki Dr, Calgary, Alberta, T3B 6A8, Canada.


| | | | | | 0 | 1 | 2 | 3 | 4 | 5 | 6 | 7 | 8 | 9 |
|---|---|---|---|---|---|---|---|---|---|---|---|---|---|---|
| **Demographics / General** | | | | | | | | | | | | | | |
| **Demographics**<br><br>See Table S2 below for further details | **Youth Demographics (Self Report)*** (Brener et al., 2024; Chin et al., 2022; DeChants et al., 2021; Potter et al., 2022; Stover et al., 2010). | Home | NA | ~7-11 items | | | | | | | | | | |
| | **Youth Demographics (Caregiver report)*** (Brener et al., 2024; Chin et al., 2022; DeChants et al., 2021; Institut de la statistique du Québec, 2024; Stover et al., 2010) | Home | NA | ~ 21 items | C | C | C | C | C | C | C | C | C | C |
| | **Caregiver & Family Demographics*** (Brener et al., 2024; Chin et al., 2022; DeChants et al., 2021; Stover et al., 2010) | Home | NA | ~ 21 items | C | C | C | C | C | C | C | C | C | C |
| **Handedness** | **Edinburgh Handedness Inventory – Short Form*** (Oldfield, 1971; Veale, 2014) | On-site | NA | 4 items | | | | | | | | | | |
| | **Adapted / Observational Edinburgh Handedness* Inventory** (Oldfield, 1971; Somerville et al., 2018); Assessed annually for 3-6yr olds; assessed at T1 only for 7-10 yr olds | On-Site | NA | 9 items / 10 mins | | | | R | R | R | R | R | R | R |
| **Physical Health & Development** | | | | | | | | | | | | | | |
| **Developmental history** (pregnancy and birth complications) | Questions adapted from **Developmental History Questionnaire*** (Barch et al., 2018; Kessler et al., 2009b, 2009a; Merikangas et al., 2009) and **IMAGEN Pregnancy and Birth Questionnaire** (Schumann et al., 2010). Assesses pregnancy conditions, substance use during second/third trimester, birth complications, and developmental milestones. | Home | NA | ~ 104 items / 15 mins | C | C | C | C | C | C | C | C | C | C |
| **Medical History** | **Medical History Questionnaire** (Barch et al., 2018; Todd et al., 2003)* | On-site | Lifetime; past year service use | ~ 77 items / 10 mins | C | C | C | C | C | C | C | C | C | C |
| **Family Psychiatric History** | Adapted from Q1K Family Psychiatric History | On-site | Lifetime | 38+ (~5mins) | C | C | C | C | C | C | C | C | C | C |
| **Concussion** | **Modified Ohio State University Traumatic Brain Injury Screen** (Bogner et al., 2017; Corrigan & Bogner, 2007) * | Home | Lifetime; Since last visit | 4+ / 5 mins | C | C | C | C | C | C | C | C | C | C |
| | **Health & Behavior Inventory** (HBI) (Ayr et al., 2009; Janusz et al., 2012) * | Home (C) / On-site (Y) | Past week; Since last visit | 20 items / 5 mins | | | | | | | | M | M | M |
| **Exercise** | **Exercise questions adapted from YRBS** (CDC, 2013)* | Home | Past 7 days | 2 items / 1 min | | | | | | | | | | |
| **Pubertal Development** | **Pubertal Development Scale (PDS)** (Petersen et al., 1988) | Home (C) / On-site (Y) | NA | ~7 / 5 mins | | | | | | | | | C | C |
| ***Sleep*** | | | | | | | | | | | | | | |

| | | | | | | | | | | | | | | |
|---|---|---|---|---|---|---|---|---|---|---|---|---|---|---|
| **Sleep Disturbances** (Bevans et al., 2019; Forrest et al., 2018) | **PROMIS Early Childhood Parent Report Short Form v1.0 – Sleep Problems – Disturbance – Short Form 4a** | Home | Past 7 days | 4 items / 1 min | | C | C | C | C | C | | | | |
| | **PROMIS Parent Proxy Short Form v1.0 – Sleep Disturbance 4a** | Home | Past 7 days | 4 items / 1 min | | | | | | | C | C | C | C |
| | **PROMIS Pediatric Short Form v1.0 – Sleep Disturbance 4a** | Home | Past 7 days | 4 items / 1 min | | | | | | | | | | |
| **Sleep Impairments** (Bevans et al., 2019; Forrest et al., 2018) | **PROMIS Early Childhood Parent Report Short Form v1.0 – Sleep Daytime Impairment 4a*** | Home | Past 7 days | 4 items / 1 min | | C | C | C | C | C | | | | |
| | **PROMIS Parent Proxy Short Form v1.0 – Sleep-Related Impairment 4** | Home | Past 7 days | 4 items / 1 min | | | | | | | C | C | C | C |
| | **PROMIS Pediatric Short Form v1.0 – Sleep-Related Impairment 4** | Home | Past 7 days | 4 items / 1 min | | | | | | | | | | |
| **Mental Health & Substance Use** | | | | | | | | | | | | | | |
| **Dimensional Psychopathology** | **Child Behavior Checklist (CBCL)** (Achenbach, 2009) | On-site | Past 6 months | 15-30 mins | | | C | C | C | C | C | C | C | C |
| | **Youth Self-Report (YSR)** (Achenbach, 2009) | On-site | Past 6 months | 15-30 mins | | | | | | | | | | |
| | **Preschool-Aged Strengths and Difficulties Questionnaire (P-SDQ)** (Goodman, 1997, 2001) *Only administered in Montreal* | Home | Past 6 months | 25 items / 5 mins | | | C | C | C | | | | | |
| | **Strengths and Difficulties Questionnaire (SDQ)** (Goodman, 1997, 2001; Goodman et al., 2000) *Only administered in Montreal* | Home | Past 6 months | 25 items / 5 mins | | | | | | C | C | C | C | C |
| **Eating** | **Ottawa Disordered Eating Screen for Youth (ODES-Y)** (Obeid et al., 2019) | Home | Past 3 months | 3 items / 2 mins | | | | | | | | | | |
| **Psychosis** | **Prodromal Questionnaire Brief (PQ-B)** (Loewy et al., 2011) | Home | Past month | 21 items / 5 mins | | | | | | | | | | |
| **Social Phobia** | **Mini-Social Phobia Inventory (Mini-SPIN)** (Connor et al., 2001; Ranta et al., 2012) | Home | Past week | 3 items / 2 mins | | | | | | | | | | |
| **Substance Use & Harms** | **CRAFFT 2.1+N** Substance Abuse Screening Test + Nicotine (Knight et al., 1999; Lee et al., 2021) | On-site | Past 12 months; lifetime harms | 0-20 items / 5 mins | | | | | | | | | | |
| | **Lifetime and past 12 month Substance Use** (recency, frequency, age of first use, non-medical prescription drug use) | On-site | Lifetime; Past 12 months | 2-5 mins | | | | | | | | | | |
| | **Alcohol Use Disorders Identification Test – Consumption (AUDIT-C)** (Bush et al., 1998; Liskola et al., 2021) | On-site | Past 12 months | 3 items / 1 mins | | | | | | | | | | |
| | **Cannabis Use Disorders Identification Test (CUDIT)** (Adamson & Sellman, 2003) | On-site | Past 12 months | 7 items / 2 mins | | | | | | | | | | |
| **Personality & Temperament** | | | | | | | | | | | | | | |

| | | | | | | | | | | | | | | |
|---|---|---|---|---|---|---|---|---|---|---|---|---|---|---|
| | Hervé, 2015) | | | | | | | | | | | | | |
| **Personality / temperament** | **Big Five Inventory-II (BFI-II; extra short form)** (Soto & John, 2017) | Home | NA | 15 / 5 mins | | | | | | | | | | |
| | **The Infant Behavior Questionnaire-Revised (IBQ-R; very short form)** (Gartstein & Rothbart, 2003; Putnam et al., n.d.; Rothbart, 1981) | Home | Past 7 days | 37 / 8 mins | C | C | | | | | | | | |
| | **Early Childhood Behavior Questionnaire (ECBQ; very short form)** (Putnam et al., 2006) | Home | Past 2 weeks | 36 / 8 mins | | | C | C | | | | | | |
| | **Children's Behavior Questionnaire (CBQ; very short form)** (Putnam & Rothbart, 2006; Rothbart et al., 2001) | Home | NA | 36 / 8 mins | | | | | C | C | C | C | | |
| **Personality traits related to substance use & psychopathology** | **Substance Use Risk Profile Scale (SURPS)** (Woicik et al., 2009) | Home / On-site | NA | 29 / 7 mins | | | | | | | | | Y | Y |
| **Cognition, Language & Motor** | | | | | | | | | | | | | | |
| **Cognition, Language & Motor** | **Bayley Scales of Infant and Toddler Development, Fourth Edition** (Bayley & Aylward, 2021) | On-site | NA | 15-60 mins | R | R | R | R | | | | | | |
| **IQ, Working Memory, Processing Speed, Verbal Comprehension, Fluid Reasoning, Visual Spatial**<br><br>See Table S3 for subtests and composite scores | **Wechsler Preschool and Primary Scale of Intelligence, Fourth Edition: Canadian** (Wechsler, 2012, 2013) | On-site | NA | 45-90 mins | | | | R | R | R | R | R | | |
| | **Wechsler Intelligence Scale for Children, Fifth Edition: Canadian** (Wechsler, 2015a, 2015b) | On-site | NA | 45-90 mins | | | | | | | | R | R | R |
| | **Wechsler Adult Intelligence Scale, Fourth Edition: Canadian** (Wechsler, 2008) | On-site | NA | 45-90 mins | | | | | | | | | | |
| **Cognitive Flexibility** | **Dimensional Change Card Sort (DCCS)** (Zelazo, 2006; Zelazo et al., 2013) | On-site | NA | 15 mins | | | | | R | R | R | R | R | R |
| **Inhibitory Control / Response Inhibition** | **Flanker Inhibitory Control** (Eriksen & Eriksen, 1974; Fan et al., 2002; Rueda et al., 2004; Zelazo et al., 2013) | On-site | NA | | | | | | R | R | R | R | R | R |
| | **Bear/Dragon** (Kochanska et al., 1996; Reed et al., 1984) | On-site | NA | 10 mins | | | R | R | R | | | | | |
| **Social & Environmental Factors** | | | | | | | | | | | | | | |
| **Digital Media Use** | Questions adapted from **Screen Time Survey / SCREENS-Q** (Barch et al., 2018; Klakk et al., 2020; Sharif et al., 2010)* | Home | Past month | 18-23 items / 5 mins | C | C | C | C | C | C | C | C | C | C |
| **Adverse Life Events & Experiences** | **PhenX Adverse Life Events** (Grant et al., 2004; Tiet et al., 2001)* | On-site | Past 12 months | 25 items / 5 mins | | | | | | | | | | |
| | **Adverse Childhood Experiences Questionnaire** (Felitti et al., 1998)* | On-site | Past 12 months | 10 items / 3 mins | C | C | C | C | C | C | C | C | C | C |
| **Caregiver self-report** | | | | | | | | | | | | | | |
| **Caregiver Digital Media Use** | Questions adapted from **Screen Time Survey** (Barch et al., 2018; Klakk et al., 2020; Sharif et al., 2010)* | Home | Past month | | C | C | C | C | C | C | C | C | C | C |
| **Caregiver Personality** | **Big Five Inventory-II (BFI-II; extra short form)** (Soto & John, 2017) | Home | NA | 15 / 5 mins | C | C | C | C | C | C | C | C | C | C |

| | | | | | | | | | | | | | | |
|---|---|---|---|---|---|---|---|---|---|---|---|---|---|---|
| **Caregiver Substance Use** | **Alcohol Use Disorder Identification Test** (Conigrave et al., 1995; Saunders et al., 1993) | Home | Past year | 10 items / 2 mins | C | C | C | C | C | C | C | C | C | C |
| | **Drug Use Disorder Identification Test** (Berman et al., 2005; Hildebrand, 2015) | Home | Past year | 10 items / 2 mins | C | C | C | C | C | C | C | C | C | C |
| **Caregiver Mental Health** | **Kessler Psychological Distress Scale – 6 items (K6)** | Home | Past month | 6 items / 1 min | C | C | C | C | C | C | C | C | C | C |

Note. * indicates questionnaires translated from English to French; ^ indicates questionnaires translated from French to English.

#### Table S2. Baseline Demographics

| Domain | Measure | Citations |
|---|---|---|
| **Youth (age 11+ years)** | | |
| School grade / level of education | PhenX<br><br>Modified to align with Canadian school systems | (Stover et al., 2010) |
| Sex assigned at birth, gender identity and sexual orientation | Adapted from PhenX and 2023 Youth Risk Behavior Survey.<br><br>Modified to align with guidelines for measuring sex and gender in youth, including intersex populations | (Brener et al., 2024; Chin et al., 2022; DeChants et al., 2021; Stover et al., 2010) |
| Romantic relationship history | Adapted from ABCD Sexual Behavior Survey | (Potter et al., 2022) |
| **Caregiver (about child)** | | |
| Child age, sex, gender identity, and ethnicity | PhenX | (Brener et al., 2024; Chin et al., 2022; DeChants et al., 2021; Stover et al., 2010) |
| Childcare | Adapted from Quebec Longitudinal Study of Child Development, 2nd edition (QLSCD 2).<br><br>Modified to include relevant options outside of Quebec | (Institut de la statistique du Québec, 2024) |
| Languages heard/spoken at home | | NA |
| Household and living arrangements | Adapted from PhenX | (Stover et al., 2010) |
| Household composition | Adapted from PhenX | (Stover et al., 2010) |
| **Caregiver (about self/family)** | | |
| Caregiver/partner age, sex, gender identity, and ethnicity | Adapted from PhenX<br><br>Modified to align with guidelines for measuring sex and gender, and with Canadian guidelines for measuring ethnicity | (Brener et al., 2024; DeChants et al., 2021; Institute for Health Information et al., 2022; Stover et al., 2010) |
| Country of origin for caregiver/s and grandparents | Adapted from PhenX | (Stover et al., 2010) |
| Caregiver education, employment and income | Adapted from PhenX<br><br>Modified response options for marital status, education level and income to align with Canadian Census | (Stover et al., 2010) |

### Table S3. Wechsler Scale Subtests Order of Administration and Composite Scores

| | Composite Scores | | | | | | | |
|---|---|---|---|---|---|---|---|---|
| **Subtest Order** | **Verbal Comprehension** | **Visual Spatial** | **Working Memory** | **Processing Speed** | **Fluid Reasoning*** | **Perceptual Reasoning** | **GAI** | **FSIQ** |
| **WPPSI-IV 2:6-3:11** | | | | | | | | |
| Receptive Vocabulary | X | | | | | | X | X |
| Block Design | | X | | | | | X | X |
| Picture Memory | | | X | | | | | X |
| Information | X | | | | | | X | X |
| Object Assembly | | X | | | | | X | X |
| Zoo Locations | | | X | | | | | X |
| **WPPSI-IV 4:0-7:7** | | | | | | | | |
| Picture Memory | | | X | | | | | X |
| Cancellation | | | | X | | | | |
| Block Design | | X | | | | | X | X |
| Receptive Vocabulary | | | | | | | | |
| Matrix Reasoning | | | | | | | X | X |
| Bug Search | | | | X | | | | X |
| Zoo Locations | | | X | | | | | |
| Similarities | X | | | | | | X | X |
| Information | X | | | | | | X | X |
| Object Assembly | | X | | | | | | |
| **WISC-V** | | | | | | | | |
| Digit Span | | | X | | | | | X |
| Coding | | | | X | | | | X |
| Block Design | | X | | | | | X | X |
| Vocabulary | X | | | | | | X | X |
| Matrix Reasoning | | | | | X | | X | X |

| | | | | | | | | |
|---|---|---|---|---|---|---|---|---|
| **Picture Span** | | | X | | | | | |
| **Symbol Search** | | | | X | | | | |
| **Similarities** | X | | | | | | X | X |
| **Figure Weights** | | | | | X | | X | X |
| **Visual Puzzles** | | X | | | | | | |
| **WAIS-IV** | | | | | | | | |
| **Digit Span** | | | X | | | | | X |
| **Coding** | | | | X | | | | |
| **Block Design** | | X | | | | X | X | X |
| **Vocabulary** | X | | | | | | X | X |
| **Matrix Reasoning** | | | | | | X | X | X |
| **Arithmetic** | | | X | | | | | X |
| **Symbol Search** | | | | X | | | | |
| **Similarities** | X | | | | | | X | |
| **Visual Puzzles** | | X | | | | X | | X |
| **Information** | X | | | | | | X | X |
| **Figure Weights** | | | | | | X | X | X |

Note. * A valid fluid reasoning composite score can only be derived from the WISC-V. The Matrix Reasoning subtest may be used as a proxy for participants who completed the WPPSI-IV or WAIS-IV
